# Viral load dynamics of SARS-CoV-2 Delta and Omicron variants following multiple vaccine doses and previous infection

**DOI:** 10.1101/2022.03.20.22272549

**Authors:** Yonatan Woodbridge, Sharon Amit, Amit Huppert, Naama M. Kopelman

## Abstract

An important, and often neglected, aspect of vaccine effectiveness is its impact on pathogen transmissibility, harboring major implications for public health policies. As viral load is a prominent factor affecting infectivity, its laboratory surrogate, qRT-PCR cycle threshold (Ct), can be used to investigate the infectivity-related component of vaccine effectiveness. While vaccine waning has previously been observed for viral load, during the Delta wave, it is yet unknown how Omicron viral load is affected by vaccination status, and whether vaccine-derived and natural infection protection are sustainable. By analyzing results of more than 460,000 individuals we show that while recent vaccination reduces Omicron viral load, its effect wanes rapidly. In contrast, a significantly slower waning rate is demonstrated for recovered COVID-19 individuals. Thus, while the vaccine is effective in decreasing morbidity and mortality, their relative minute effect on transmissibility and rapid waning call for reassessment of the scientific justification for “vaccine certificate”, as it may promote false reassurance and promiscuous behavior.

---

Vaccine effectiveness (VE) is usually measured as the protection from infection or from clinical disease [1,2]. However, this definition neglects an important aspect - the potential risk of transmission given infection. The latter has major implications on devising public health policies to reduce the spread of the pathogen. Viral load (VL) is the most prominent factor affecting infectivity and is negatively correlated with the cycle threshold (Ct) values of quantitative real-time polymerase chain reaction (qRT-PCR). Thus Ct values of routine diagnostic SARS-CoV-2 PCR tests are a readily available surrogate allowing clinicians and researchers to estimate infectiousness [3,4].

In Israel, the BNT162b2 vaccination campaign was launched on December 19th, 2020. In light of a resurgence caused by the Delta variant, a third dose (“booster”) campaign was launched on July 29th, 2021, for individuals who had received the second dose at least 5 months earlier. Currently, about 6.1 million people received two doses and ∼4.4 million people received three doses [5]. Nonetheless, Israel, like the rest of the world, is experiencing an unprecedented Omicron resurgence. In order to protect the elderly population, a fourth dose was administered to people over 60, high-risk individuals, and healthcare workers, starting from January 3rd 2022. Consequently, more than 700,000 people have received four doses[5].

In this retrospective study, we combined the meticulated national vaccination data with Ct data of four laboratories performing SARS-CoV-2 PCR tests. Studies on the Delta variant have shown that vaccinated individuals have lower VL, thus considered less infective, and that this effect wanes as time elapses [6,7]. We augment these studies by examining the influence and temporal dynamics of vaccination and recovery on Omicron VL and how this changes with time since vaccination/infection for the Omicron variant using nationwide PCR data. Notably, the waning effect of infection-induced protection has not been thoroughly analyzed before in terms of VL and infectivity. We further compare presumed infectivity of individuals vaccinated with 2,3, or 4 doses versus COVID-19 recovered individuals.

Ct data of positive tests were obtained from four molecular labs, two of which are major Israeli Health Maintenance Organization labs, representing together approximately 40% of the Isareli population, and the other two are labs commissioned to perform tests for the Israeli Ministry of Health (MoH). All of the PCR tests included in this study were part of MoH surveillance scheme, and charge free for the consumer. We analyzed Ct values dating June 15, 2021 to January 29, 2022, divided into two periods of Delta and Omicron dominance (see Extended Data). Separate analyses were conducted for the viral nucleocapsid gene (N, 441,748 measurements) and envelope gene (E, 328,865 measurements). The patterns observed for E were similar to those of N (see Extended Data). To circumvent platform and other methodological variances between laboratories each lab dataset was analyzed both separately and combined, with similar patterns observed (see Extended Data).

We first performed multivariate linear regression analysis on Ct values of each variant with vaccination status, laboratory, age, sex and calendar time (7-days bins) as covariates (Extended Data Table 2A). Vaccination status was defined as unvaccinated, 2-dose (divided to 3 bins, 10-39 days, 40-69, >70 days post-vaccination), 3-dose (divided to 3 bins, 10-39, 40-69, >70 days), 4-dose, or recovered; Due to the small number of individuals in the 2-dose (early) groups for the Omicron period, all the 2-dose groups were combined. Fig.1 depicts the adjusted Ct values for both periods (see Extended Data Figure 2A for extended results on each lab).

**Fig 1.**
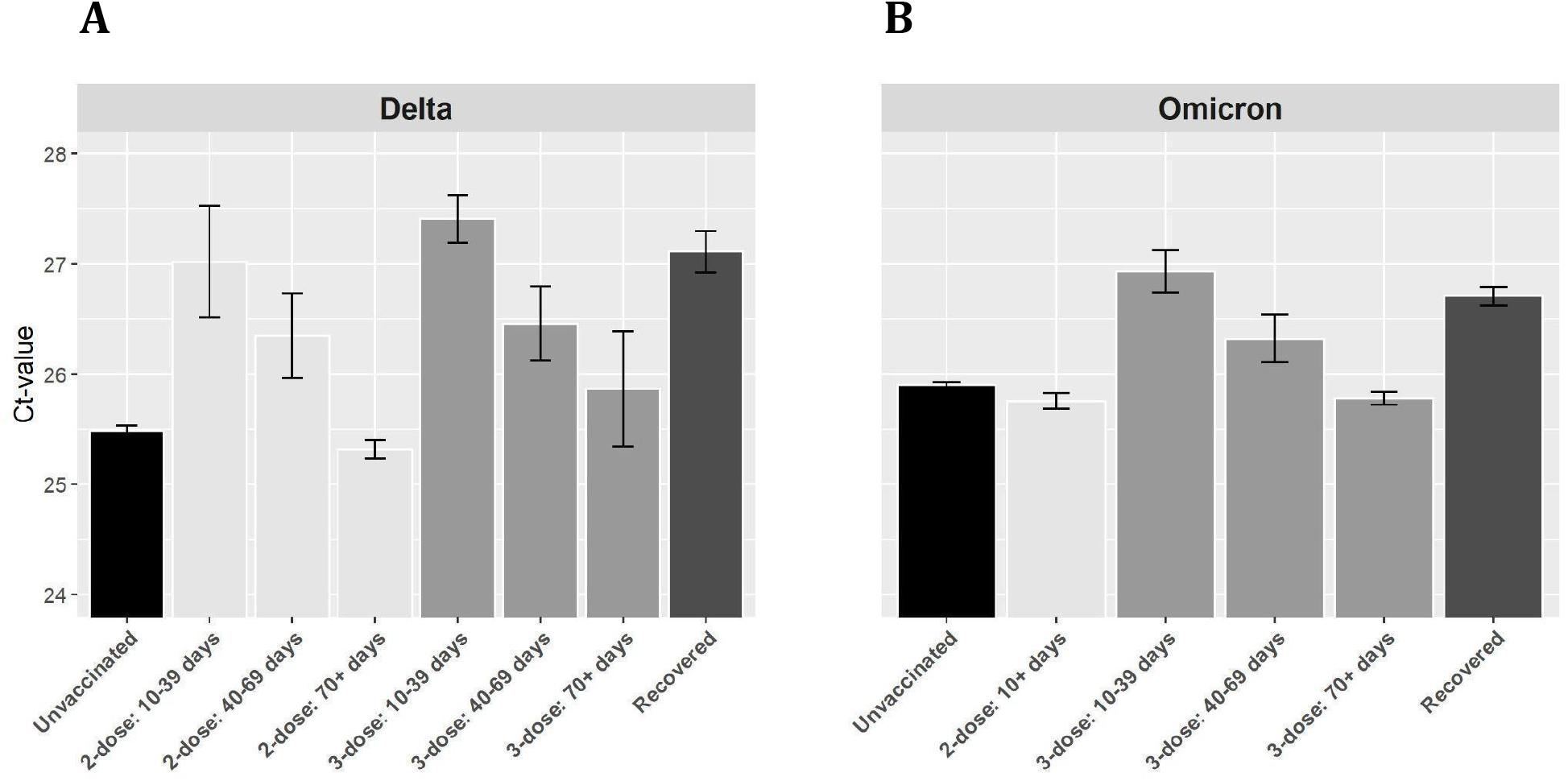
Ct values of the gene N. Adjusted Ct values of different vaccination statuses, measured by four laboratories, for the Delta **(A)** and Omicron **(B)** variants. Means were obtained from the weighted sum of age, sex and calendar time, and the intercepts. Error bars represent 95% CI’s around the means, obtained by using the estimated distribution of all four labs together (see Materials and Methods).

Like previously reported [7], during the Delta surge, 2-dose noticeably decreases VL (Fig. 1A). For the 2nd-dose early cohort (10-39 days), mean Ct is about 1.3 Ct units higher than that of unvaccinated, corresponding to more than two-fold decrease in VL; However, this protection wanes rapidly as time elapses since vaccination, and VL reaches a level similar to that of the unvaccinated by day 70. For the 3-dose (early) cohort, VL is even lower than for the 2-dose (early) cohort, but once again rapid waning follows, and by day 70 VL reaches the baseline level of the unvaccinated. Notably, VL of the recovered cohort is similar to that of the 2-dose (early) and 3-dose (early) cohorts.

During the Omicron period (Fig. 1B), only a recent 3rd-dose decreases VL among vaccinees, and is similar to infection-derived protection. Otherwise, the differences in VL for the unvaccinated, 2- and late 3-dose groups (Ct values of 25.9, 25.7 and 25.7 respectively) are negligible. In general, the effect of immune status for Omicron is less pronounced than from Delta even upon recent receipt of the 3rd vaccine dose or infection, as manifested by a reduced Ct-values gaps between these groups and the unvaccinated. The relative difference between the recently vaccinated (3-dose, 10-39 days) and the unvaccinated is smaller in Omicron (1.03, 95% CI 0.84-1.22) compared to Delta (1.92, 95% CI 1.71-2.13). Similarly, the relative difference between recovered and unvaccinated is 1.63 (95% CI 1.45-1.81)in Delta, while in Omicron it is reduced to 0.8 (95% CI, 0.72-0.88). These gaps are reduced by a two-fold in Omicron, possibly due to host immune waning and viral evasion [9] (see Extended Data Table 2A for full regression results).

Since the 4-dose jab was administered mainly to the elderly (60+, 90.9% of all 4-dose receivers), we conducted a separate analysis for this age group, pooling the 2- and 3-dose subgroups together. The results, presented in Fig. 2, reveal that shortly after receiving the 4th dose, VL of the vaccinated individuals (ages >60) reaches levels similar yet slightly higher than those of recovered individuals from the same age group.

**Fig 2.**
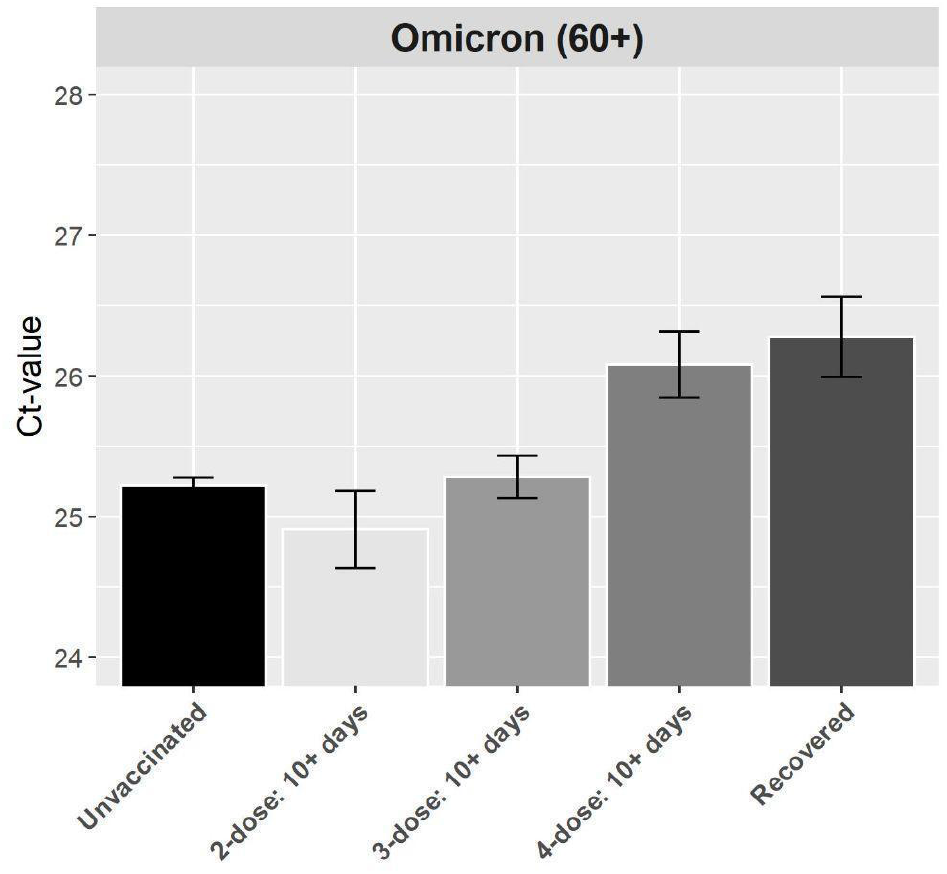
Ct values of the gene N for the elderly population during Omicron. Adjusted Ct values of different vaccination statuses for the Omicron variants in individuals of age >60. Means were obtained from the weighted sum of age, sex and calendar time, and the intercepts. Error bars represent 95% CI’s around the means, obtained by using the estimated distribution of all labs together (see Materials and Methods).

Goldberg et al. [10] have shown that natural infection protection wanes over time, with a rate that is slower than that of vaccine-derived protection. Here we demonstrate for the first time that waning of natural infection protection pertains also to VL, for both Delta and Omicron. Fig. 3 shows a clear and consistent waning trend for the recovered, for both Delta (Fig. 3A) and Omicron (Fig. 3B) variants. In contrast to the rapid waning observed for vaccinated individuals (Fig. 1&2), VL of the previously infected does not reach the baseline level of the unvaccinated even after extended periods of time (>12 months) and for individuals originally infected with the Wuhan or Alpha strains (Fig. 3).

**Fig 3.**
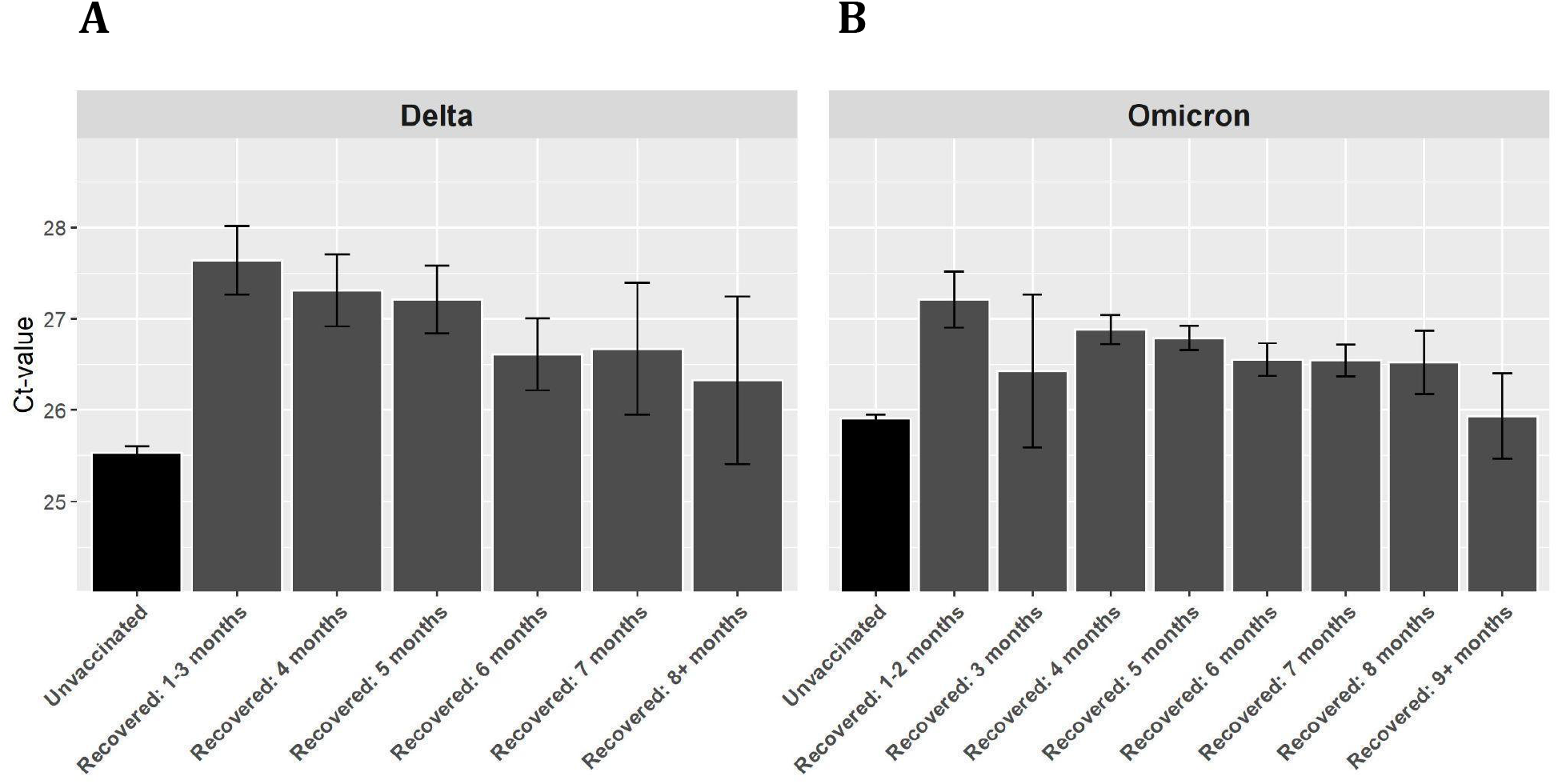
Ct values of the gene N for unvaccinated and recovered individuals. Ct-values for unvaccianted and recovered patients, partitioned by number of months after recovery, for the Delta **(A)** and Omicron **(B)** variants. Means were obtained from the weighted sum of age, sex and calendar time, and the intercepts. Error bars represent 95% CI’s around the means, obtained by using the estimated distribution of all four labs together (see Materials and Methods).

Some precaution should be taken interpreting Ct data, in light of lab-specific standards [11] and clinical correlations. Regression is a standard approach which partially adjusts for such differences. In addition, Ct values may contain temporal biases due to changing policies and health-seeking behaviors. The inclusion of calendric date as an explanatory variable in the regressions(see Extended Data Tables 1A & 2A) (following [10]), as well as considering only the first positive test for each patient, mitigate this possible bias. Additionally, we tested the robustness of the results by examining the patterns also for the gene E (Extended Data Fig. 3A), conducting separate single lab regressions (Extended Data Fig. 4A), accounting for temporal biases using R0 instead of calendar time (Extended Data Table 4A, Analysis 1), and narrowing the period for Delta as well as restricting age ranges for both variants (Extended Data Table 4A, Analysis 2).

This study indicates that overall the presumed vaccination-related immunity to SARS-CoV-2 has only a negligible long term (>70-days) effect on Ct value, a common surrogate for VL and infectiousness. The combination of vaccine waning and vaccine evasion are most likely the drivers of this finding. In lieu of several prominent publications describing vaccine effectiveness in prevention morbidity and hospitalization for Omicron [12,13], this study questions the role of current vaccination campaigns in harnessing the transmissibility of COVID-19 at a time scale greater than two months. Likewise, the scientific justification for “vaccination certificates” should be reassessed, taking into consideration the short term effects in reducing VL, as it may contribute to false reassurance and promiscuous behavior, and prove counterproductive as an epidemiologic restriction measure. Further studies should assess the differential benefits of SARS-CoV-2 vaccines in alleviating disease vs. preventing pathogen spread. Should this gap prove consistent, it may have major ramifications on global pandemic preparedness, vaccination rollout and medical inequity as well as other public health measures.

## Data Availability

All data produced in the present study are available upon reasonable request to the authors

## Acknowledgments

This work was supported by the Israel Science Foundation KillCorona-Curbing Coronavirus Research Program (grant no. 3663/19 to N. M. K.). The funders had no role in study design, data collection and analysis, decision to publish or preparation of the manuscript. The authors would like to thank Arnona Ziv, Micha Mandel, Yair Goldberg and Ronen Fluss.

The study was approved by an Institutional Review Board (IRB) of the Sheba Medical Center. Helsinki approval number: SMC-8228-21.

## Materials and Methods

### Dataset construction

We used a nationwide database of Ct values from positive cases, measured by four different laboratories. Samples were collected between June 15, 2021 to Jan 29, 2022. The dataset contained over four million records of positive PCR tests with Ct-values. Records contained Ct measurements for the genes N, E, Orf1ab or S genes. Results are presented for N and E, two genes whose measurements make up ∼90% of all available data for the four laboratories. Ct values<10 or >40 units were removed from the dataset, since such values are likely the result of reading errors. A negligible number of records such samples were identified and removed from the four labs, N & E final dataset (9 records).

Multiple Ct measurements for the same individual and gene may belong to the same or different infection event. Sequences of Ct measurements within a single 90 days interval were defined as belonging to the same infection events. For each such sequence, only the first (earliest) Ct value was taken. Multiple infection events for a single individual were included if the time difference between the last measurement of the first sequence and the first measurement of the second sequence was at least 90 days. For the second infection, the patient’s status was defined as ‘recovered’.

Overall, our analyses, performed on the data of four labs, contained more than 460,000 individuals, 441,748 Ct measurements for the gene N, and 328,865 measurements for the gene E. Using encrypted patients’ identity numbers, we merged Ct data with demographic information and vaccination data, to determine the patient’s age, sex, and vaccination status. The merged data contained both Ct date and PCR sampling date. For the Ct measurements included in this study, the number of days between these two dates was at most a single day. Since PCR date is the actual sampling date, these dates were used for analyses.

Vaccination statuses were determined for each patient and infection event, based on the PCR date. The following definitions were used to group individuals:

– *Unvaccinated*: Including up to 6 days after the first dose.
– *2-dose 10-39*: From 10 days after the second dose up to 39 days after the second dose.
– *2-dose 40-69*: From 40 days after the second dose up to 69 days after the second dose.
– *2-dose 70+*: From 70 days onwards after the second dose.
– *3-dose 10-39*: From 10 days after the third dose up to 39 days after the third dose.
– *3-dose 40-69*: From 40 days after the third dose up to 69 days after the third dose.
– *3-dose 70+*: From 70 days onwards after the third dose.
– *4-dose 10+*: From 10 days onwards after the fourth dose.
– *Recovered*: Individuals who had a previous infection event with a positive PCR test at least 90 days prior to the current infection. This group includes unvaccinated and vaccinated recovered individuals, excluding individuals that received more than a single shot after recovery. The latter were excluded since the Israeli MoH policy was to allow only one dose for recovered individuals.

Patients who had the infection between first and second dose (i.e, from 7 day after first dose until 10 days after second dose) were not included in the analysis.

We divided the follow-up study into two separate periods, each dominated by a different variant:

– *Delta time period*: June 15 to Dec 1 2021 (>90% of the cases identified as Delta, see [14]).
– *Omicron time period*: Dec 28 2021 to Jan 29 2022 (>90% of the cases identified as Omicron, see [14]).

These periods are also in accordance with the first documented case of Omicron infection in Israel (see Extended Data Figure 1A).

To account for temporal effects in our regression analyses, we partitioned the Delta and Omicron time periods into 7-day time intervals (bins), using PCR dates to classify Ct measurements. Age groups were defined as

0-11, 12-15, 16-39, 40-59, and 60 or older. Due to national policy, individuals of age 0-11 were not present in most cohorts (they were not vaccinated until recently), and this group was thus excluded from the main analysis. Nonetheless, this age group was included in parts of the sensitivity analysis presented in the appendix.

### Statistical analysis

Our main tools in assessing the effect of different factors on Ct-values are the linear and quantile regression. On examination of the different cohorts, we used cohort, age category, sex, and categorized calendar date as explanatory variables. Daily Ct values may have been sampled from patients at different stages of the infection (i.e, time from infection). We thus also examined the median and a lower quartile of Ct values, controlling for age-of-infection variability (see Extended Data Tables 2A & 3A). To compute error bars in Figs. 1-3, as well as Extended Data Figs. 2A-4A, we used the estimated regression coefficients mean and variance, and through the multivariate normal distribution calculated the (0.025,0.975)-percentiles to obtain confidence intervals.

## Extended data tables

**Table 1A.** Number of samples, mean (s.d.) and median of all N-gene Ct values, grouped by laboratory, cohort and gene, for Delta and Omicron time periods. Only ages above 11 were included in this table.

| Lab | Cohort | Delta |  |  | Omicron |  |  |
| --- | --- | --- | --- | --- | --- | --- | --- |
|  |  | N | Mean(s.d) | Median | N | Mean(s.d) | Median |
| Lab1 | unvaccinated | 13271 | 25.4 (5.04) | 24.71 | 19986 | 26.07 (4.74) | 25.65 |
|  | 2-dose: 10-39 | 141 | 27.57 (4.87) | 26.91 | 170 | 25.72 (4.45) | 25.25 |
|  | 2-dose: 40-69 | 223 | 26.37 (4.77) | 25.87 | 142 | 26.74 (4.84) | 26.81 |
|  | 2-dose: 70+ | 15247 | 25.36 (5.01) | 24.48 | 11230 | 26.03 (4.78) | 25.59 |
|  | 3-dose: 10-39 | 657 | 28.19 (4.59) | 28.39 | 1225 | 27.11 (4.58) | 26.83 |
|  | 3-dose: 40-69 | 252 | 26.8 (4.6) | 26.52 | 703 | 26.53 (4.72) | 26.1 |
|  | 3-dose: 70+ | 113 | 27.08 (4.27) | 26.76 | 44585 | 25.94 (4.74) | 25.49 |
|  | 4-dose |  |  |  | 1554 | 25.73 (4.6) | 25.5 |
|  | Recovered (+/- vaccine) | 733 | 27.67 (5.01) | 27.79 | 6895 | 27.16 (4.58) | 26.95 |
| Lab2 | unvaccinated | 9027 | 26.61 (5.65) | 25.9 | 706 | 26.42 (4.17) | 26.34 |
|  | 2-dose: 10-39 | 57 | 27.08 (5.72) | 25.95 | 1 | 26.34 () | 26.34 |
|  | 2-dose: 40-69 | 105 | 27.64 (5.69) | 26.45 | 4 | 26.63 (5.18) | 27.67 |
|  | 2-dose: 70+ | 6634 | 26.22 (5.51) | 25.26 | 342 | 25.93 (4.52) | 25.79 |
|  | 3-dose: 10-39 | 340 | 27.91 (5.48) | 27.25 | 6 | 29.09 (1.96) | 29.13 |
|  | 3-dose: 40-69 | 135 | 28.38 (5.82) | 28 | 15 | 27.13 (4.22) | 26 |
|  | 3-dose: 70+ | 64 | 26.66 (5.79) | 25.77 | 665 | 25.5 (4.5) | 25.21 |
|  | 4-dose |  |  |  | 1 | 34.2 () | 34.2 |
|  | Recovered (+/- vaccine) | 936 | 28.06 (5.66) | 27.62 | 390 | 26.47 (4.23) | 25.84 |
| Lab3 | unvaccinated | 13784 | 25 (4.61) | 24.61 | 16210 | 25.87 (4.01) | 25.8 |
|  | 2-dose: 10-39 | 90 | 26.52 (4.18) | 25.98 | 85 | 25.41 (3.8) | 25.59 |
|  | 2-dose: 40-69 | 144 | 25.91 (4.32) | 25.45 | 87 | 25.69 (3.58) | 25.22 |
|  | 2-dose: 70+ | 15060 | 24.74 (4.54) | 24.19 | 10189 | 25.64 (3.98) | 25.5 |
|  | 3-dose: 10-39 | 555 | 26.57 (4.36) | 26.34 | 628 | 26.96 (3.84) | 27.06 |
|  | 3-dose: 40-69 | 155 | 26.63 (4.37) | 26.3 | 645 | 26.2 (3.96) | 26.1 |
|  | 3-dose: 70+ | 68 | 26.23 (4.69) | 26.18 | 26750 | 25.79 (4.03) | 25.64 |
|  | 4-dose |  |  |  | 732 | 26.36 (4.24) | 26.42 |
|  | Recovered (+/- vaccine) | 579 | 26.33 (4.65) | 26.29 | 4989 | 26.37 (3.96) | 26.31 |
| Lab4 | unvaccinated | 11580 | 25.51 (4.63) | 25.03 | 8816 | 25.64 (4.4) | 25.97 |
|  | 2-dose: 10-39 | 79 | 26.57 (4.68) | 26.03 | 59 | 25.62 (4.8) | 25.03 |
|  | 2-dose: 40-69 | 168 | 25.8 (4.5) | 25.03 | 44 | 27.26 (4.68) | 27.5 |
|  | 2-dose: 70+ | 11623 | 25.24 (4.52) | 25.03 | 4924 | 25.46 (4.41) | 25.03 |
|  | 3-dose: 10-39 | 716 | 26.8 (4.42) | 27.03 | 367 | 26.15 (4.53) | 26.03 |
|  | 3-dose: 40-69 | 353 | 26.49 (4.58) | 26.03 | 370 | 26.26 (4.44) | 26.03 |
|  | 3-dose: 70+ | 145 | 27.08 (4.58) | 27.03 | 19943 | 25.37 (4.46) | 25.03 |
|  | 4-dose |  |  |  | 222 | 24.82 (4.32) | 25.03 |
|  | Recovered (+/- vaccine) | 762 | 26.81 (4.21) | 27.03 | 3340 | 26.52 (4.32) | 26.03 |

### Regression results for the gene N

**Table 2A.**
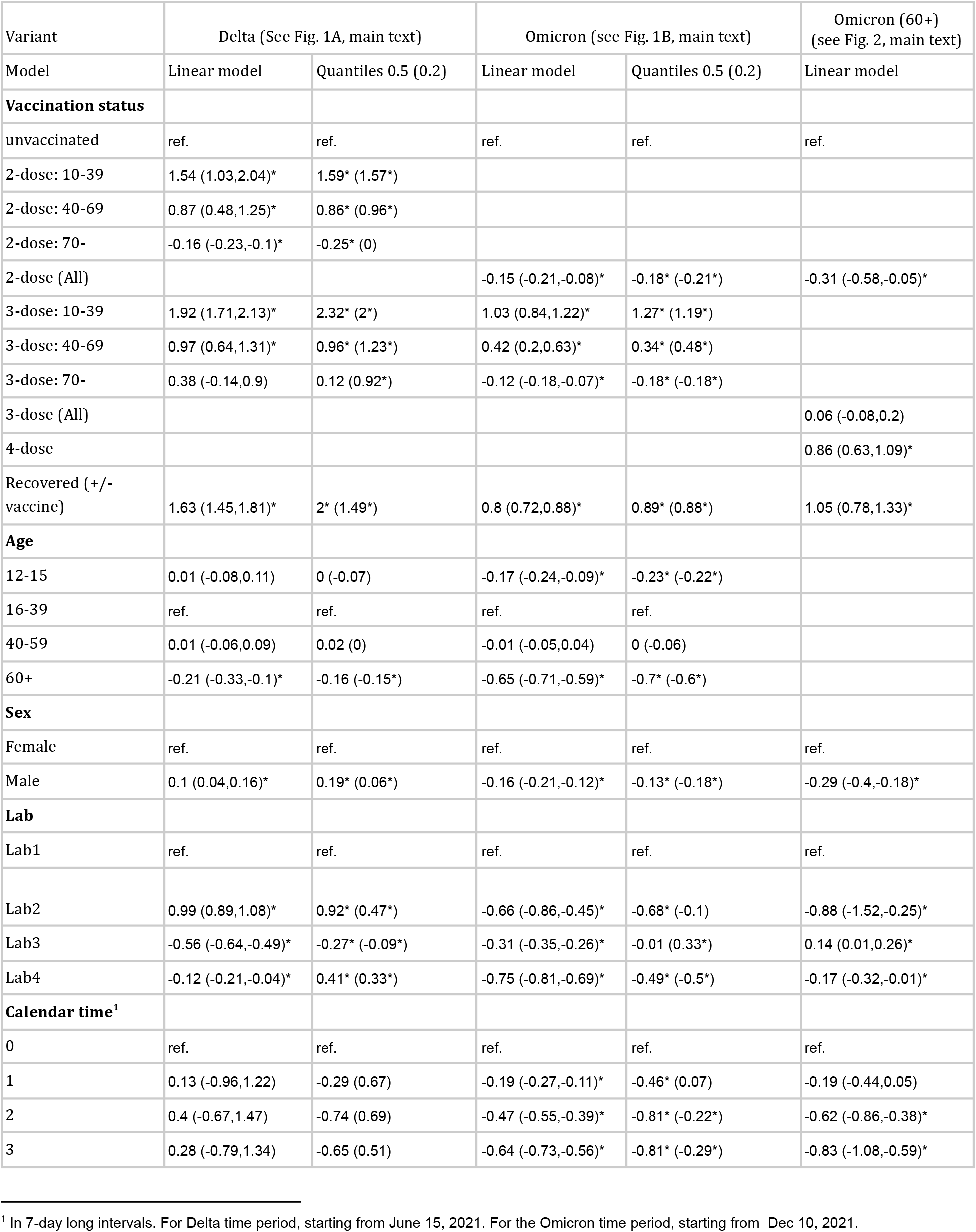
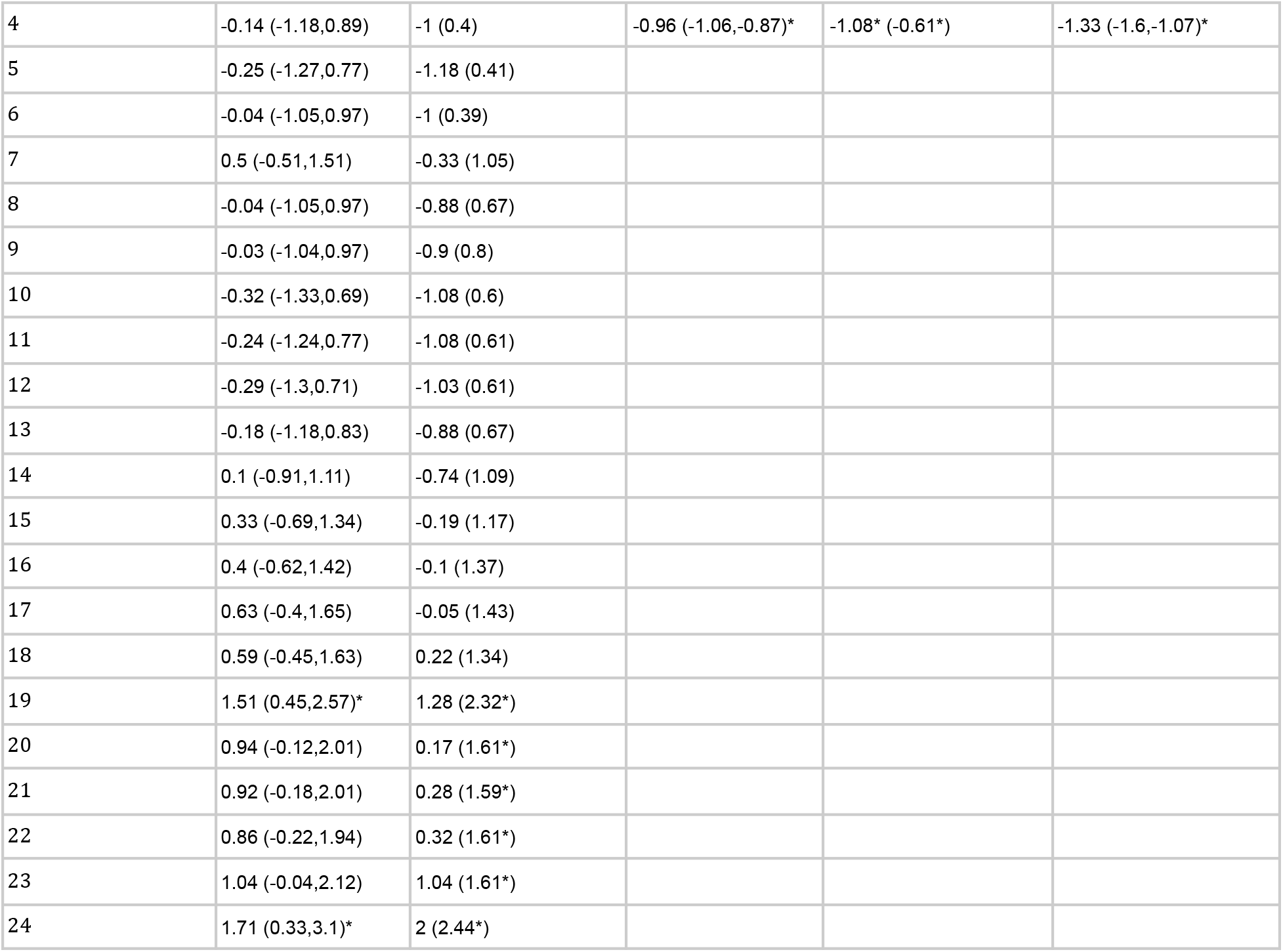
Regression analyses’ results for Ct values. The linear regressions are the primary analyses discussed in the text and presented in Figures 1-3, while the quantile regressions were conducted as secondary analyses. The revealed trends are highly similar between the linear and quantile analyses. 95% CI are given in parentheses. For quantile regression, median and 0.2 quartile (in parentheses) are provided. (*) denote 0.05 significance.

| Variant | Delta (See Fig. 1A, main text) |  | Omicron (see Fig. 1B, main text) |  | Omicron (60+)<br>(see Fig. 2, main text) |
| --- | --- | --- | --- | --- | --- |
| Model | Linear model | Quantiles 0.5 (0.2) | Linear model | Quantiles 0.5 (0.2) | Linear model |
| **Vaccination status** |  |  |  |  |  |
| unvaccinated | ref. | ref. | ref. | ref. | ref. |
| 2-dose: 10-39 | 1.54 (1.03,2.04)* | 1.59* (1.57*) |  |  |  |
| 2-dose: 40-69 | 0.87 (0.48,1.25)* | 0.86* (0.96*) |  |  |  |
| 2-dose: 70- | -0.16 (-0.23,-0.1)* | -0.25* (0) |  |  |  |
| 2-dose (All) |  |  | -0.15 (-0.21,-0.08)* | -0.18* (-0.21*) | -0.31 (-0.58,-0.05)* |
| 3-dose: 10-39 | 1.92 (1.71,2.13)* | 2.32* (2*) | 1.03 (0.84,1.22)* | 1.27* (1.19*) |  |
| 3-dose: 40-69 | 0.97 (0.64,1.31)* | 0.96* (1.23*) | 0.42 (0.2,0.63)* | 0.34* (0.48*) |  |
| 3-dose: 70- | 0.38 (-0.14,0.9) | 0.12 (0.92*) | -0.12 (-0.18,-0.07)* | -0.18* (-0.18*) |  |
| 3-dose (All) |  |  |  |  | 0.06 (-0.08,0.2) |
| 4-dose |  |  |  |  | 0.86 (0.63,1.09)* |
| Recovered (+/- vaccine) | 1.63 (1.45,1.81)* | 2* (1.49*) | 0.8 (0.72,0.88)* | 0.89* (0.88*) | 1.05 (0.78,1.33)* |
| **Age** |  |  |  |  |  |
| 12-15 | 0.01 (-0.08,0.11) | 0 (-0.07) | -0.17 (-0.24,-0.09)* | -0.23* (-0.22*) |  |
| 16-39 | ref. | ref. | ref. | ref. |  |
| 40-59 | 0.01 (-0.06,0.09) | 0.02 (0) | -0.01 (-0.05,0.04) | 0 (-0.06) |  |
| 60+ | -0.21 (-0.33,-0.1)* | -0.16 (-0.15*) | -0.65 (-0.71,-0.59)* | -0.7* (-0.6*) |  |
| **Sex** |  |  |  |  |  |
| Female | ref. | ref. | ref. | ref. | ref. |
| Male | 0.1 (0.04,0.16)* | 0.19* (0.06*) | -0.16 (-0.21,-0.12)* | -0.13* (-0.18*) | -0.29 (-0.4,-0.18)* |
| **Lab** |  |  |  |  |  |
| Lab1 | ref. | ref. | ref. | ref. | ref. |
| Lab2 | 0.99 (0.89,1.08)* | 0.92* (0.47*) | -0.66 (-0.86,-0.45)* | -0.68* (-0.1) | -0.88 (-1.52,-0.25)* |
| Lab3 | -0.56 (-0.64,-0.49)* | -0.27* (-0.09*) | -0.31 (-0.35,-0.26)* | -0.01 (0.33*) | 0.14 (0.01,0.26)* |
| Lab4 | -0.12 (-0.21,-0.04)* | 0.41* (0.33*) | -0.75 (-0.81,-0.69)* | -0.49* (-0.5*) | -0.17 (-0.32,-0.01)* |
| **Calendar time<sup>1</sup>** |  |  |  |  |  |
| 0 | ref. | ref. | ref. | ref. | ref. |
| 1 | 0.13 (-0.96,1.22) | -0.29 (0.67) | -0.19 (-0.27,-0.11)* | -0.46* (0.07) | -0.19 (-0.44,0.05) |
| 2 | 0.4 (-0.67,1.47) | -0.74 (0.69) | -0.47 (-0.55,-0.39)* | -0.81* (-0.22*) | -0.62 (-0.86,-0.38)* |
| 3 | 0.28 (-0.79,1.34) | -0.65 (0.51) | -0.64 (-0.73,-0.56)* | -0.81* (-0.29*) | -0.83 (-1.08,-0.59)* |

<sup>1</sup> In 7-day long intervals. For Delta time period, starting from June 15, 2021. For the Omicron time period, starting from Dec 10, 2021.

Table 2A. Regression analyses' results for Ct values.
|  |  |  |  |  |  |
| --- | --- | --- | --- | --- | --- |
| 4 | -0.14 (-1.18,0.89) | -1 (0.4) | -0.96 (-1.06,-0.87)* | -1.08* (-0.61*) | -1.33 (-1.6,-1.07)* |
| 5 | -0.25 (-1.27,0.77) | -1.18 (0.41) |  |  |  |
| 6 | -0.04 (-1.05,0.97) | -1 (0.39) |  |  |  |
| 7 | 0.5 (-0.51,1.51) | -0.33 (1.05) |  |  |  |
| 8 | -0.04 (-1.05,0.97) | -0.88 (0.67) |  |  |  |
| 9 | -0.03 (-1.04,0.97) | -0.9 (0.8) |  |  |  |
| 10 | -0.32 (-1.33,0.69) | -1.08 (0.6) |  |  |  |
| 11 | -0.24 (-1.24,0.77) | -1.08 (0.61) |  |  |  |
| 12 | -0.29 (-1.3,0.71) | -1.03 (0.61) |  |  |  |
| 13 | -0.18 (-1.18,0.83) | -0.88 (0.67) |  |  |  |
| 14 | 0.1 (-0.91,1.11) | -0.74 (1.09) |  |  |  |
| 15 | 0.33 (-0.69,1.34) | -0.19 (1.17) |  |  |  |
| 16 | 0.4 (-0.62,1.42) | -0.1 (1.37) |  |  |  |
| 17 | 0.63 (-0.4,1.65) | -0.05 (1.43) |  |  |  |
| 18 | 0.59 (-0.45,1.63) | 0.22 (1.34) |  |  |  |
| 19 | 1.51 (0.45,2.57)* | 1.28 (2.32*) |  |  |  |
| 20 | 0.94 (-0.12,2.01) | 0.17 (1.61*) |  |  |  |
| 21 | 0.92 (-0.18,2.01) | 0.28 (1.59*) |  |  |  |
| 22 | 0.86 (-0.22,1.94) | 0.32 (1.61*) |  |  |  |
| 23 | 1.04 (-0.04,2.12) | 1.04 (1.61*) |  |  |  |
| 24 | 1.71 (0.33,3.1)* | 2 (2.44*) |  |  |  |

### Regression results for the gene E

**Table 3A.**
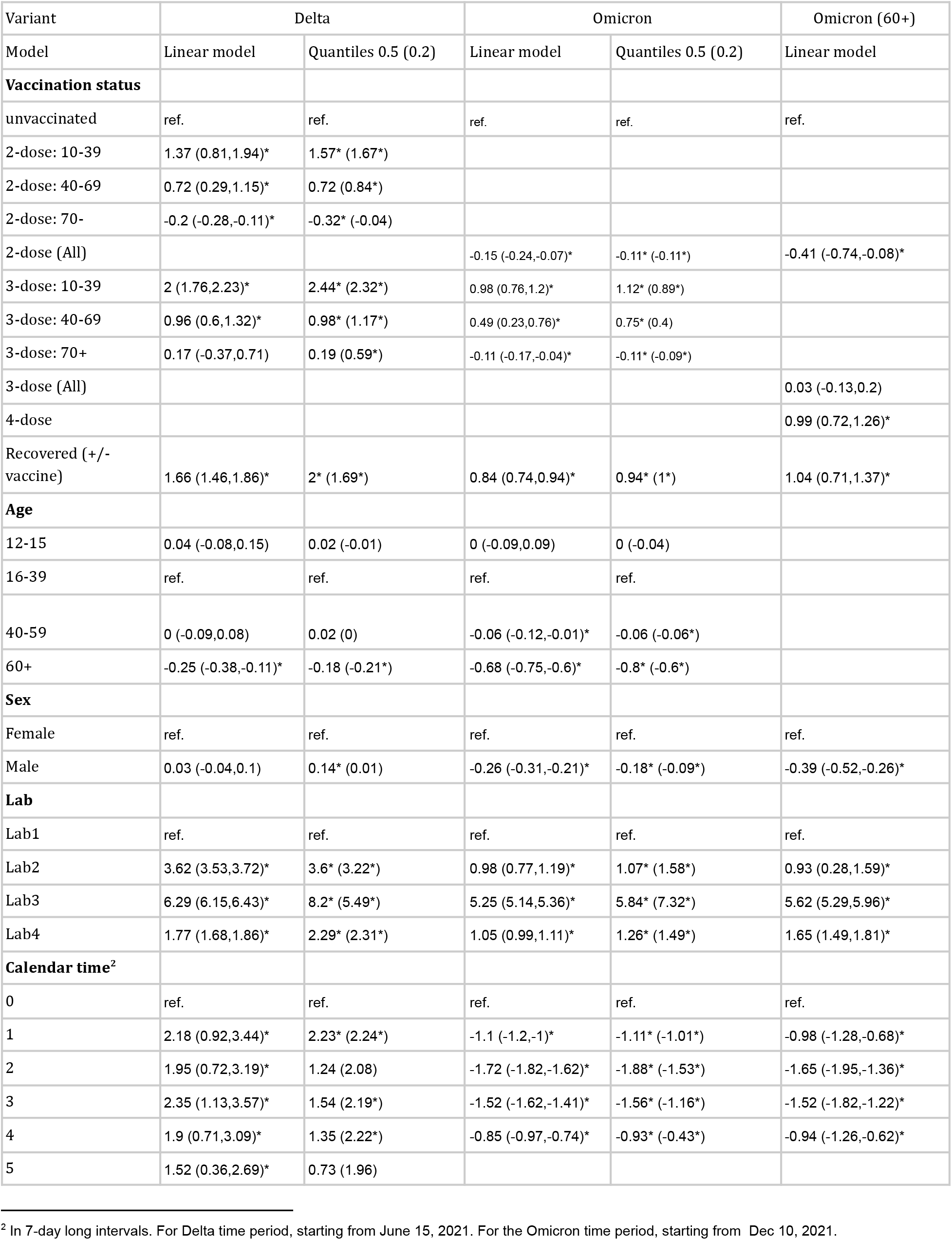
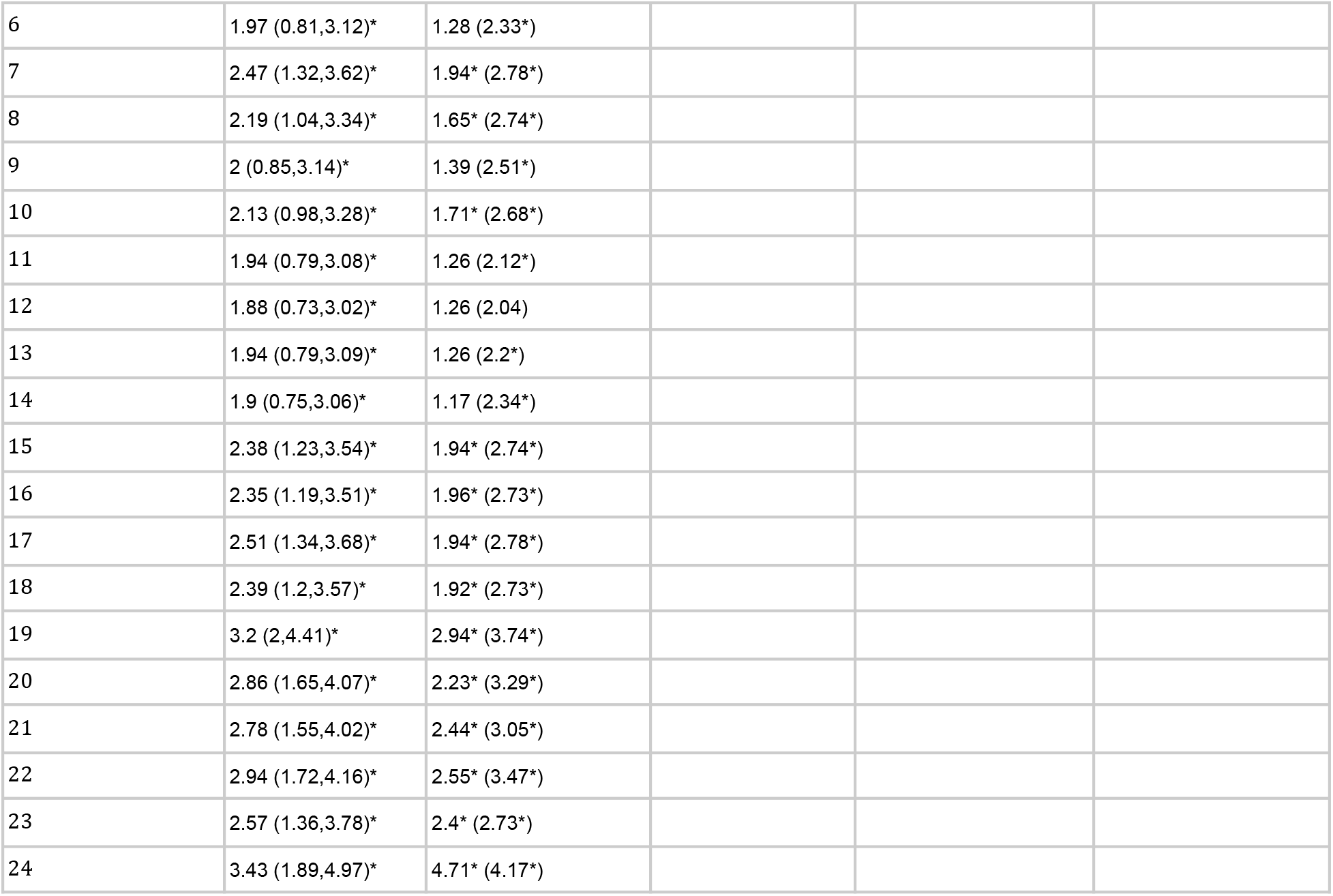
Extended regression results from Table 2A for E-gene Ct-values, together with calendar time coefficients. Linear and quantile regression results on Ct-value, estimated separately for Delta and Omicron time periods. 95% CI are given in parentheses. For quantile regression, median and 0.2 quartile (in parentheses) are provided. (*) denote 0.05 significance.

| Variant | Delta |  | Omicron |  | Omicron (60+) |
| --- | --- | --- | --- | --- | --- |
| Model | Linear model | Quantiles 0.5 (0.2) | Linear model | Quantiles 0.5 (0.2) | Linear model |
| **Vaccination status** |  |  |  |  |  |
| unvaccinated | ref. | ref. | ref. | ref. | ref. |
| 2-dose: 10-39 | 1.37 (0.81,1.94)* | 1.57* (1.67*) |  |  |  |
| 2-dose: 40-69 | 0.72 (0.29,1.15)* | 0.72 (0.84*) |  |  |  |
| 2-dose: 70- | -0.2 (-0.28,-0.11)* | -0.32* (-0.04) |  |  |  |
| 2-dose (All) |  |  | -0.15 (-0.24,-0.07)* | -0.11* (-0.11*) | -0.41 (-0.74,-0.08)* |
| 3-dose: 10-39 | 2 (1.76,2.23)* | 2.44* (2.32*) | 0.98 (0.76,1.2)* | 1.12* (0.89*) |  |
| 3-dose: 40-69 | 0.96 (0.6,1.32)* | 0.98* (1.17*) | 0.49 (0.23,0.76)* | 0.75* (0.4) |  |
| 3-dose: 70+ | 0.17 (-0.37,0.71) | 0.19 (0.59*) | -0.11 (-0.17,-0.04)* | -0.11* (-0.09*) |  |
| 3-dose (All) |  |  |  |  | 0.03 (-0.13,0.2) |
| 4-dose |  |  |  |  | 0.99 (0.72,1.26)* |
| Recovered (+/- vaccine) | 1.66 (1.46,1.86)* | 2* (1.69*) | 0.84 (0.74,0.94)* | 0.94* (1*) | 1.04 (0.71,1.37)* |
| **Age** |  |  |  |  |  |
| 12-15 | 0.04 (-0.08,0.15) | 0.02 (-0.01) | 0 (-0.09,0.09) | 0 (-0.04) |  |
| 16-39 | ref. | ref. | ref. | ref. |  |
| 40-59 | 0 (-0.09,0.08) | 0.02 (0) | -0.06 (-0.12,-0.01)* | -0.06 (-0.06*) |  |
| 60+ | -0.25 (-0.38,-0.11)* | -0.18 (-0.21*) | -0.68 (-0.75,-0.6)* | -0.8* (-0.6*) |  |
| **Sex** |  |  |  |  |  |
| Female | ref. | ref. | ref. | ref. | ref. |
| Male | 0.03 (-0.04,0.1) | 0.14* (0.01) | -0.26 (-0.31,-0.21)* | -0.18* (-0.09*) | -0.39 (-0.52,-0.26)* |
| **Lab** |  |  |  |  |  |
| Lab1 | ref. | ref. | ref. | ref. | ref. |
| Lab2 | 3.62 (3.53,3.72)* | 3.6* (3.22*) | 0.98 (0.77,1.19)* | 1.07* (1.58*) | 0.93 (0.28,1.59)* |
| Lab3 | 6.29 (6.15,6.43)* | 8.2* (5.49*) | 5.25 (5.14,5.36)* | 5.84* (7.32*) | 5.62 (5.29,5.96)* |
| Lab4 | 1.77 (1.68,1.86)* | 2.29* (2.31*) | 1.05 (0.99,1.11)* | 1.26* (1.49*) | 1.65 (1.49,1.81)* |
| **Calendar time<sup>2</sup>** |  |  |  |  |  |
| 0 | ref. | ref. | ref. | ref. | ref. |
| 1 | 2.18 (0.92,3.44)* | 2.23* (2.24*) | -1.1 (-1.2,-1)* | -1.11* (-1.01*) | -0.98 (-1.28,-0.68)* |
| 2 | 1.95 (0.72,3.19)* | 1.24 (2.08) | -1.72 (-1.82,-1.62)* | -1.88* (-1.53*) | -1.65 (-1.95,-1.36)* |
| 3 | 2.35 (1.13,3.57)* | 1.54 (2.19*) | -1.52 (-1.62,-1.41)* | -1.56* (-1.16*) | -1.52 (-1.82,-1.22)* |
| 4 | 1.9 (0.71,3.09)* | 1.35 (2.22*) | -0.85 (-0.97,-0.74)* | -0.93* (-0.43*) | -0.94 (-1.26,-0.62)* |
| 5 | 1.52 (0.36,2.69)* | 0.73 (1.96) |  |  |  |

<sup>2</sup> In 7-day long intervals. For Delta time period, starting from June 15, 2021. For the Omicron time period, starting from Dec 10, 2021.

Table 3A. Extended regression results from Table 2A for E-gene Ct-values, together with calendar time coefficients.
|  |  |  |
| --- | --- | --- |
| 6 | 1.97 (0.81,3.12)* | 1.28 (2.33*) |
| 7 | 2.47 (1.32,3.62)* | 1.94* (2.78*) |
| 8 | 2.19 (1.04,3.34)* | 1.65* (2.74*) |
| 9 | 2 (0.85,3.14)* | 1.39 (2.51*) |
| 10 | 2.13 (0.98,3.28)* | 1.71* (2.68*) |
| 11 | 1.94 (0.79,3.08)* | 1.26 (2.12*) |
| 12 | 1.88 (0.73,3.02)* | 1.26 (2.04) |
| 13 | 1.94 (0.79,3.09)* | 1.26 (2.2*) |
| 14 | 1.9 (0.75,3.06)* | 1.17 (2.34*) |
| 15 | 2.38 (1.23,3.54)* | 1.94* (2.74*) |
| 16 | 2.35 (1.19,3.51)* | 1.96* (2.73*) |
| 17 | 2.51 (1.34,3.68)* | 1.94* (2.78*) |
| 18 | 2.39 (1.2,3.57)* | 1.92* (2.73*) |
| 19 | 3.2 (2,4.41)* | 2.94* (3.74*) |
| 20 | 2.86 (1.65,4.07)* | 2.23* (3.29*) |
| 21 | 2.78 (1.55,4.02)* | 2.44* (3.05*) |
| 22 | 2.94 (1.72,4.16)* | 2.55* (3.47*) |
| 23 | 2.57 (1.36,3.78)* | 2.4* (2.73*) |
| 24 | 3.43 (1.89,4.97)* | 4.71* (4.17*) |

## Extended data figures

**Fig 1A.**
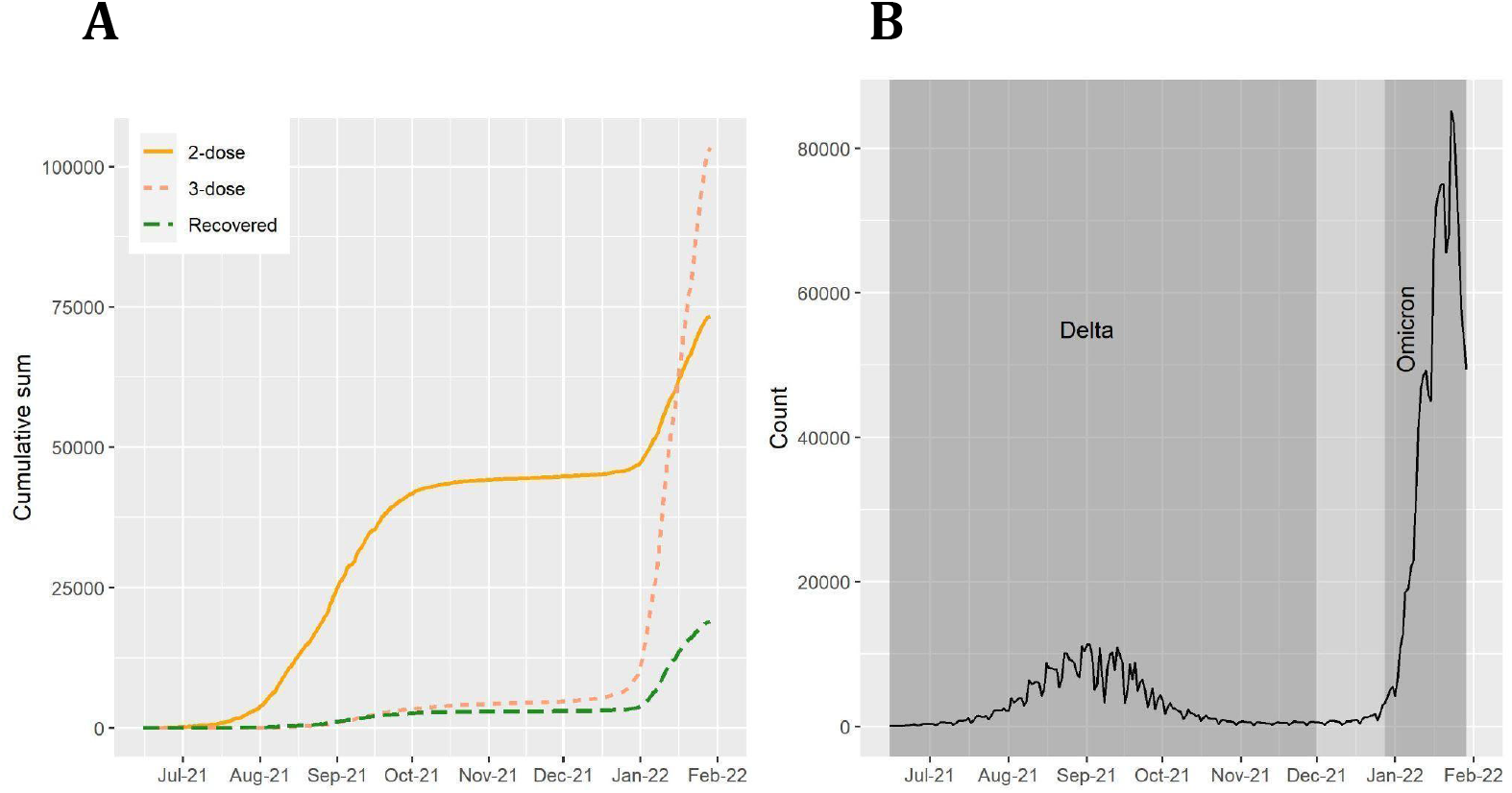
**A**. The cumulative number of N-gene Ct measurements by date, for 2-does, 3-dose, and recovered individuals, for a combined dataset of all four labs, from June 15, 2021until Jan 29, 2022. **B**. Number of positive cases, as reported by the Israeli MOH. Shaded areas highlight the Delta and Omicron time periods, as defined in the main text.

**Fig 2A.**
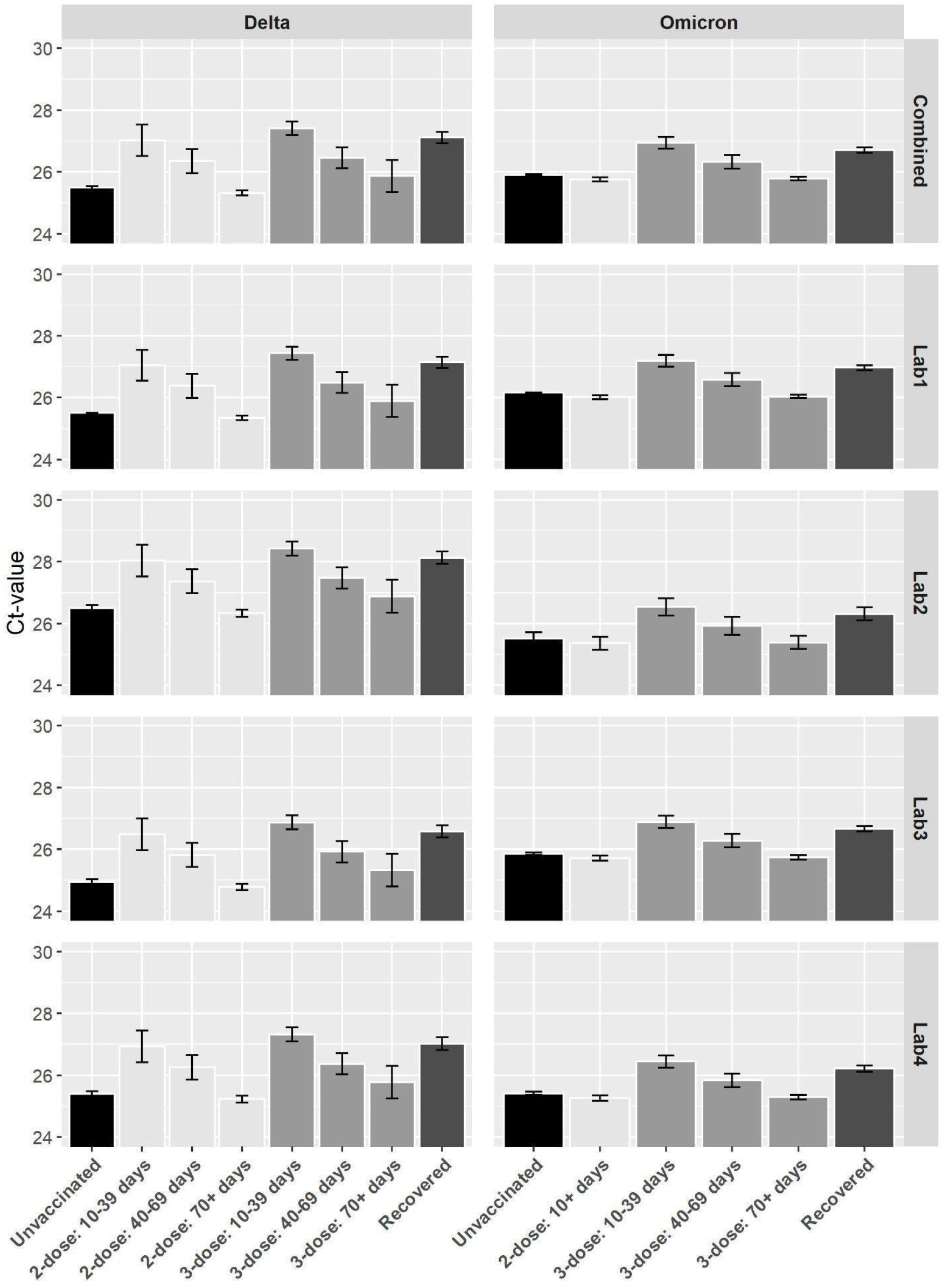
**N-gene Ct values** for different vaccination statuses, measured by four laboratories and combined, for the Delta and Omicron variants (left and right panels, respectively), using the multivariate regression coefficients (see Table 2A). Results are provided for each lab in a separate row, note the similar pattern in all labs. Error bars represent 95% CI’s around the means. Means were obtained from the weighted sum of age, sex and calendar time (using their frequency for each variant), together with the intercept and the corresponding cohort and lab for each column bar. CI’s were obtained using the estimated distribution of each pair of cohort-lab coefficients. As to “Combined”, CI’s were obtained from the estimated distribution of all labs together with each of the cohorts coefficients.

**Fig 3A.**
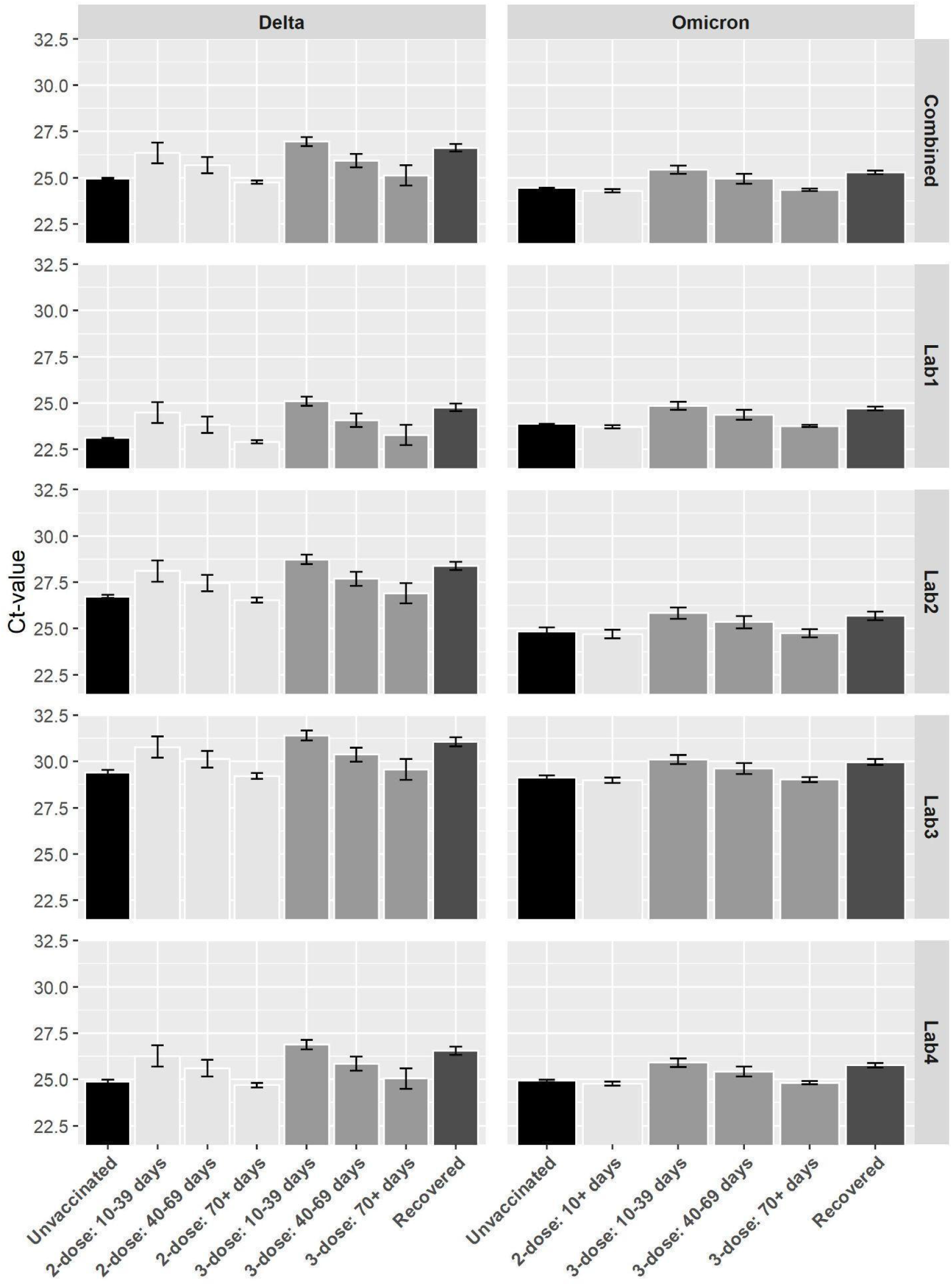
**E-gene Ct values** for different vaccination statuses, measured by four laboratories and combined, for the Delta and Omicron variants (left and right panels, respectively), using the multivariate regression coefficients. Note that both E and N have similar patterns. Results are provided for each lab in a separate row, note the similar pattern in all labs. Error bars represent 95% CI’s around the means. Means were obtained from the weighted sum of age, sex and calendar time (using their frequency for each variant), together with the intercept and the corresponding cohort and lab for each column bar. CI’s were obtained using the estimated distribution of each pair of cohort-lab coefficients. As to “Combined”, CI’s were obtained from the estimated distribution of all labs together with each of the cohorts coefficients.

## Appendix I

### Sensitivity Analysis

Fig. 4A presents the results of regression analyses performed separately on each lab, in order to examine each lab independently, thus accounting for lab variability. The consistency of the results supports our assumption that vaccination status affects Ct values in a similar manner across all labs, regardless of different lab procedures and measurement standards.

**Fig 4A.**
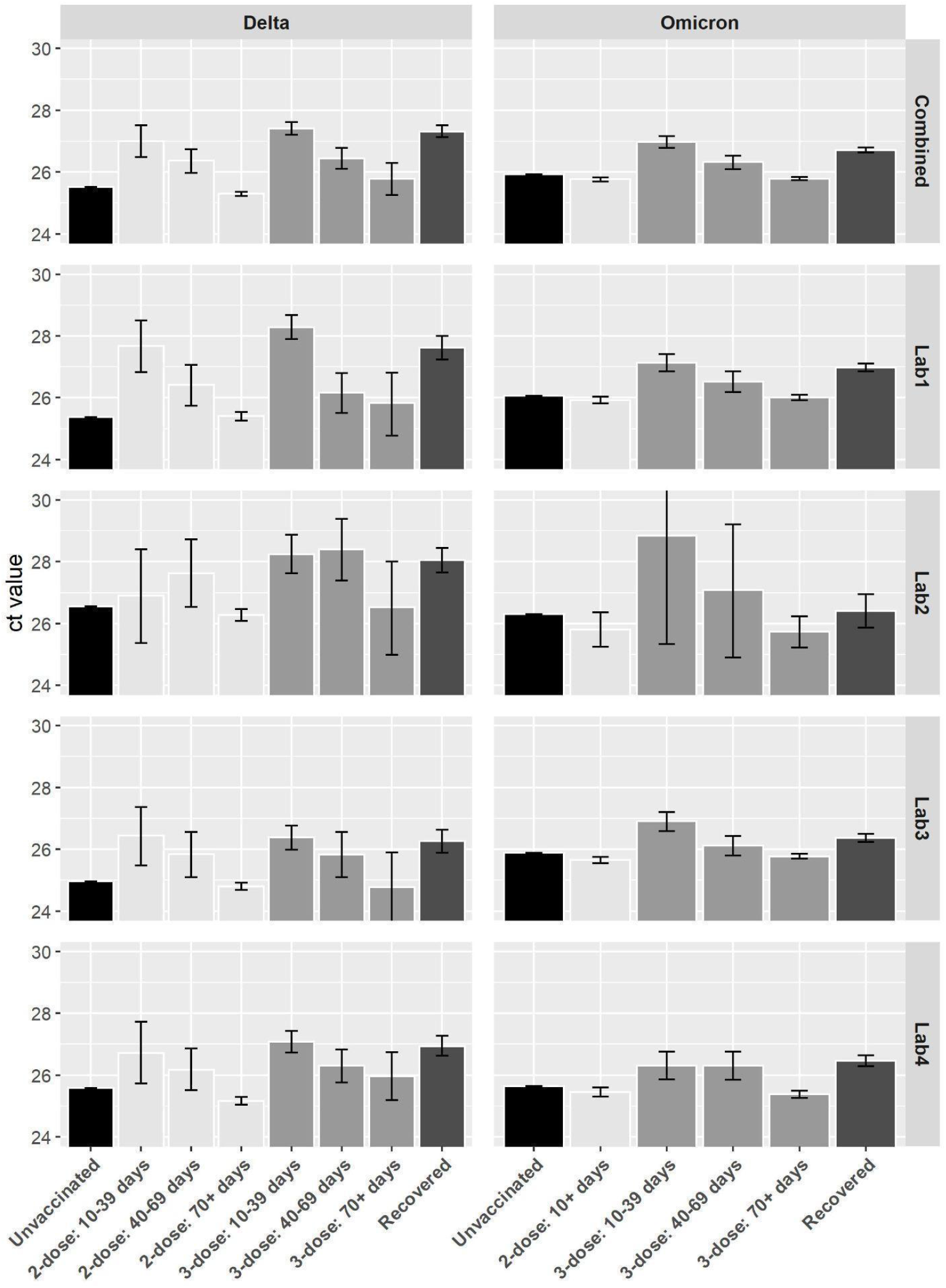
**N-gene Ct values** for different vaccination cohorts, for the Delta and Omicron variants based on regression analyses performed separately for each lab.. Error bars represent 95% CI’s around the means. Means were obtained from the weighted sum of age, sex and calendar time (using their frequency for each pair of lab-variant), together with the intercept and the corresponding cohort for each column bar. As to ‘Combined’, means were obtained from the weighted sum of age, sex, calendar time and lab, together with the intercept and the corresponding cohort for each column bar. CI’s were obtained using the estimated distribution of each of the cohort coefficients.

Table 4A presents two parts of a sensitivity analysis. In Analysis 1, we performed linear regression with the same covariates as in Fig. 1 & Table 2A, but replaced calendar time with the R0 variable, thus accounting for temporal effects in a different manner. We observe similar patterns in terms of the coefficients, both for Delta and Omicron, as shown in Table 4A. In Analysis 2, we used a restricted follow-up time study for Delta (between Sep 07 and Oct 11, 2021), as well as certain ages for both variants (12-20 for Delta, 5-20 for Omicron. note that in Omicron we included ages 5-11, that were excluded from the main analyses), to mitigate temporal and age effects. Results are provided in the rightmost columns of Table 4A.

**Table 4A.**
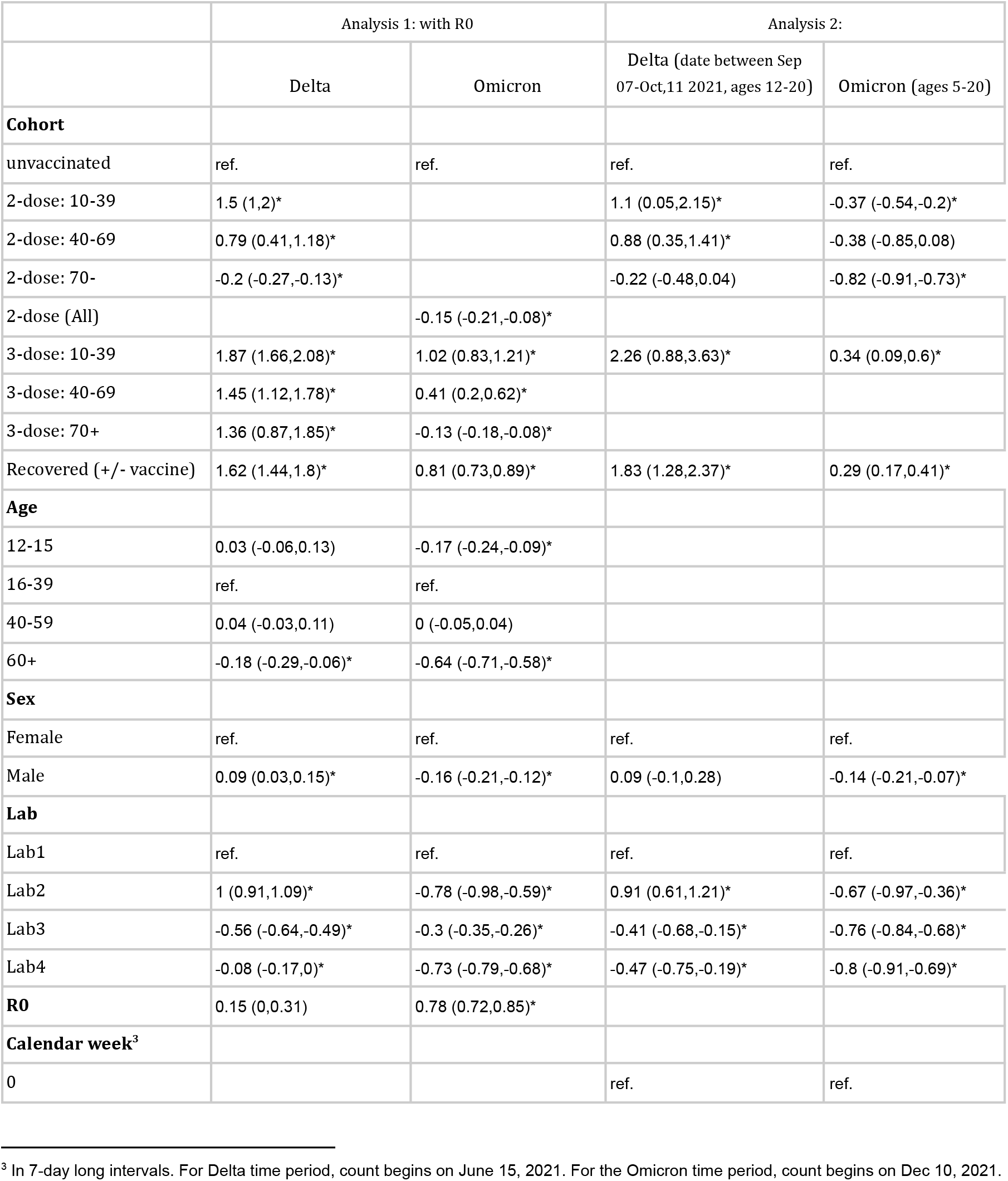
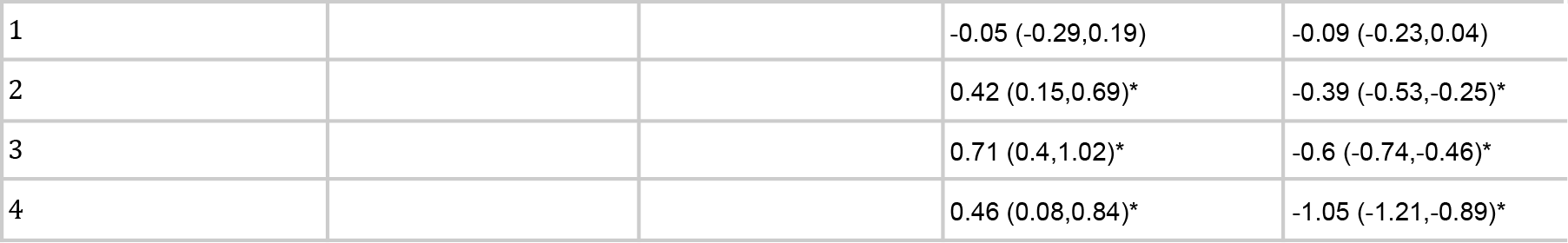
Additional regression analyses. Analysis 1 (using R0 instead of calendar time, in Delta and Omicron). Analysis 2 (truncation of follow-up study time and age in Delta, truncation of age in Omicron).

